# Sleeping with One Eye Open: Lived Experiences of Informal Caregivers Regarding Nighttime Agitation in People with Dementia

**DOI:** 10.64898/2026.03.27.26349496

**Authors:** Ajda Flisar, Maarten Van Den Bossche, Evelien Coppens, Chantal van Audenhove, Jessie Dezutter

## Abstract

Nighttime agitation (NA) is a prevalent and challenging phenomenon affecting people with dementia (PwD), often resulting in premature institutionalization. Yet, informal caregivers’ perspectives on this phenomenon remain underexplored. We conducted 15 in-depth interviews with informal caregivers to gain insight into their experiences and reactions to NA. Thematic analysis identified seven sub-themes related to carers’ experience and eight sub-themes concerning their reactions. These themes emerged across three levels, namely, PwD, informal caregiver and the environment. Most phenomena occurred at a dyadic level between PwD and informal caregiver, highlighting the potential of interventions targeting dyadic coping. Informal caregivers feel insufficiently supported when sleep disturbances co-occur with NA. They primarily rely on self-initiated strategies and learn by experience. Caregivers mention the need for more advanced knowledge and skills in reacting to co-occurrence of sleep disturbances with NA or systemic support in terms of dealing with emergencies. Caregivers also reflect extensively on the impact of challenging behaviors during the night on their mental and physical well-being. Notably, no non-pharmacological interventions for NA adequately address the themes identified in this study, highlighting the urgent need for integrative approaches and recognition of caregiver wellbeing as a core outcome, not a secondary consideration in interventions.

## Introduction

With an increasing ageing population, experts predict that the number of dementia cases is expected to triple by 2050, reaching 139 million worldwide (Dementia Statistics, Alzheimer’s Disease International, n.d.). The International Psychogeriatric Association (IPA) defines dementia as “a significant impairment in cognitive function, often associated with behavioral and psychological symptoms in dementia (BPSD)”. Various types of dementia exist, with the most common ones being Alzheimer’s disease, vascular dementia, dementia with Lewy Bodies, frontotemporal dementia, and Parkinson’s dementia. Throughout the progression of the disease, almost all individuals develop BPSD (Steinberg et al., 2008). Agitation is known to be the most common and burdensome BPSD, as it decreases the quality of life of people with dementia (PwD; Karttunen et al., 2011) and imposes a substantial strain on caregivers (Shi et al., 2018). Given the high reliance on informal caregivers in dementia care, understanding their perspectives on challenging dementia symptoms is crucial. This study examines the experiences and reactions of informal caregivers in relation to one particularly disruptive symptom, namely nighttime agitation (NA).

### Nighttime Agitation in Dementia

While dementia is most commonly associated with progressive cognitive decline, BPSD are equally critical, particularly in the context of care provision. Among these symptoms, agitation is one of the most prevalent and challenging symptoms. It typically manifests as excessive motor activity, verbal or physical aggression, and is often accompanied by emotional distress (Cummings et al., 2015). At the level of the PwD, agitation is associated with accelerated cognitive decline and disease progression (Canevelli et al., 2013; Ismail et al., 2016). Agitation very often leads to (over)prescribing psychotropic medication (Salzman et al., 2008; Wetzels et al., 2011), which is associated with dangerous side effects and increased morbidity and mortality (Ballard et al., 2009). Agitation in dementia may also occur during the night, commonly referred to as NA, although no consensus definition currently exists. Different authors use terms such as *NA*, *nocturnal restlessness*, *nightly disturbance*, and *sleep-related agitation* to describe similar phenomena (Cipriani et al., 2015; Greenblum & Rowe, 2012; McCurry et al., 2006; Rose et al., 2011; Webster et al., 2020). NA includes nocturnal behaviors encompassing wandering, verbal agitation, aggression, searching behavior and inapropriate activities for the nighttime (Rose et al., 2011). It is strongly reinforced by sleep disturbances and circadian rhythm disruptions, creating a vicious cycle in which poor sleep fuels agitation and agitation further disrupts sleep (Rose et al., 2011). Despite the significant impact on patient and caregiver wellbeing, NA remains in many studies an unaddressed symptom. At the level of an individual, NA in combination with decreased sleep quality decreases the quality of life for PwD (Tatsumi et al., 2009). At the level of society, NA poses a financial burden as it is associated with (premature) institutionalization and, in the long term, increased economic costs of long-term care (Ancoli-Israel & Vitiello, 2006; Costa et al., 2018; Hope et al., 1998; Sano et al., 2024; Schoenmekers et al., 2009). The toll of NA is, however, also reflected at the level of caregivers, who experience significant distress and decreased mental and physical well-being (Schein et al., 2022).

### Source of distress for informal caregivers

Informal caregivers are individuals without professional training, such as family members, friends, or paid helpers, who provide care at home for people with dementia (Etters et al., 2008). With dementia being an irreversible condition, informal caregivers are expected to carry an increased responsibility as the condition of the PwD declines (Chiao et al., 2015). As the word “informal” already implies, informal caregivers usually require even more support, psychoeducation and training in dealing with challenging symptoms in dementia (Chiao et al., 2015). As life expectancy continues to rise across populations, informal caregivers will be increasingly confronted with prolonged disease trajectories and a greater prevalence of dementia-related symptoms, increasing caregiver burden.

Caregiver burden, defined by Zarit et al. (1986), refers to the ‘degree to which a carer’s emotional or physical health, social life or financial status has suffered as a result of caring for their relative (page 261). This burden significantly contributes to diminished mental health, including anxiety and depression (Cooper et al., 2007), as well as physical health, such as cardiovascular disease and immune functional problems (Xu et al., 2020). In the long term, it may even contribute to increased mortality (Pristavec & Luth, 2020).

The presence of BPSD during the night can be particularly distressing for informal caregivers.

Research consistently shows that sleep disturbances and nighttime behavioral problems in PwD are strongly linked to poorer sleep quality among their caregivers (Castro et al., 2009). Disrupted sleep in caregivers, in turn, exacerbates caregiver burden and symptoms of depression (Castro et al., 2009; Creese et al., 2008; Kim et al., 2014). With NA continuously interrupting the nights of caregivers, this symptom, in many cases, forces informal caregivers to opt for (premature) institutionalization of PwD (Rose et al., 2011).

### Research gap

Despite growing evidence that NA is burdensome and more distressing for informal caregivers compared to professional carers (Tan et al., 2005), little is known about how informal caregivers *experience* these episodes and how they *respond* to them in home settings. Existing qualitative studies have examined caregivers’ strategies for managing BPSD more broadly (De Vugt et al., 2004; Huis In Het Veld et al., 2016; Leung et al., 2021; Moore et al., 2013; Polenick et al., 2020; Turner et al., 2015; Van Vracem et al., 2016). What these studies all have in common is that they highlight a limited repertoire of strategies available to informal caregivers despite experiencing a wide range of BPSD in PwD. In line with this finding, these studies show that informal caregivers mainly use self-initiated management strategies to manage changes in behavior and mood of their relative. These, for example, include encouraging activity, identifying triggers, using restraint or paternalistic interventions, and addressing physiological needs (Moore et al., 2013). These studies also found that, next to management strategies for BPSD, informal caregivers emphasize self-management strategies to maintain their own physical and mental well-being. In addition to caregivers’ experiences with BPSD in general, studies have also addressed the same question specifically for sleep disturbances in PwD. Importantly, these studies examined sleep solely, excluding NA. As such, understanding sleep disturbances has already been addressed by Gibson et al. (2014), who reflected upon caregivers’ challenges such as being woken up, not being able to fall back asleep, trips to the bathroom and daytime sleepiness. Similarly, Huisman et al. (2025) identified the main challenges related to sleep changes in PwD, namely difficulties in maintaining time orientation, disrupted day/night routine, irregularities, and concerns of informal caregivers at night and environmental factors that support or disturb sleep. The authors underscored the importance of presenting the results within the dyadic relationship, highlighting the importance of contextualizing the experiences of informal caregivers and possible support for them in relation to the functioning of the PwD.

Despite the contributions of the above-mentioned studies, only one study has examined NA in PwD. However, it was conducted in a nursing home setting and focused on care professionals’ experiences and reactions (Van Vracem et al., 2016). No study to date has explored the lived experiences of *informal* caregivers specifically in relation to NA in PwD *living at home*, nor how caregivers navigate the emotional, practical, and relational challenges of this behavior in their daily life. This gap is notable given that NA often represents a turning point for caregiver exhaustion and decisions about institutionalization. Indeed, Kim et al. (2014), for example, found that caregivers’ perceptions of NA significantly predicted caregiver burden, whereas objective data did not. This supports the idea that informal caregivers’ lived experience and perceptions offer unique insights into NA that differ from the views of professionals, and this knowledge is highly needed.

To address this gap, the present study explores the lived experiences of informal caregivers of PwD living at home regarding NA. The study furthermore examines how informal caregivers react to and manage these episodes. By focusing specifically on NA rather than on agitation in general, we aim to provide a more nuanced understanding of a phenomenon that profoundly impacts both members of the caregiving dyad. Insights of this study may contribute to a deeper understanding of NA and may inform the development of interventions that can better support informal caregivers.

## Method

### Participants

We recruited participants using purposive sampling, deliberately screening for participants who are informal caregivers of PwD experiencing NA. The inclusion criteria were: (1) older than 18 years, (2) currently providing care for a person living with dementia living at home and experiencing NA, (3) Slovenian as their mother tongue, and (4) willingness to share personal experiences of caring for a person with dementia at home. Detailed demographic and caregiving characteristics of the sample are presented in Table 1.

**Table 1.** Information about the sample.

| Pseudonym | Gender (F/M) | Providing care for | Household information |
| --- | --- | --- | --- |
| Maria | F | Mother | SH |
| Yasmin | F | Mother | DH – independent living |
| Olivia | F | Mother and father | SH |
| Anna | F | Mother and father | SH |
| Laura | F | Mother | SH |
| Stephanie | F | Mother and father-in-law | SH |
| Nina | F | Father | SH |
| Peter | M | Mother | SH |
| Alfie | M | Wife | SH |
| George | M | Mother | DH – independent living |
| Emma | F | Father-in-law | SH |
| Lucy | F | Mother-in-law | SH |
| Jane | F | Father | SH |
| Kate | F | Mother | SH |
| Iris | F | Mother | DH – assisted apartment<br>facility |

*Note F- Female, M – Male; SH – same household, DH – different household*

**Table 2.** Presentation of the themes and subthemes.

| **Experiences** of informal caregivers about NA | **Reactions** of informal caregivers to NA |
| --- | --- |
| Sleeping with one eye open | Establishing stability through routine |
| Emotional toll of stressful nights | Emotional reactivity |
| Perceived necessity of medication | Medication as the last resort |
| Fear for the safety of their loved one | Managing risk through safety solutions |
| Expecting the unexpected | Nighttime crisis management |
| When caring comes at a cost | Learning the hard way |
| External and environmental factors shaping the nights | Communication as comfort |
|  | Seeking support resources |

### Procedure

The study was preregistered on OSF (https://osf.io/nqrk2/)^1^. Participants were recruited in Slovenia through the distribution of flyers in public places such as libraries and pharmacies, as well as via social media. The study flyer was also shared online in a Facebook group for informal caregivers of PwD called “Dementia” to reach an audience from different regions in the country. Once participants reached out to us expressing their interest in participating, we called them to explain the study in more detail, answer any of their questions, and confirm that they met all inclusion criteria.

Full study information was provided to enable informed decision-making. Upon confirmation of participation, the interview date and location were scheduled in accordance with the participant’s preferences. At the time of the interview, participants received an information letter and informed consent, which they had to sign before the start of the interview. The interviews were conducted between August and October 2024 at a location with sufficient privacy (depending on the wishes of the participant) and lasted 45-90 minutes. Each participant received a gift card of 20 euros for the participation. The study was approved by the ethical commission of the University of Leuven, Belgium (SMEC code: G-2024-8112-R3(AMD)).

The first author conducted 15 in-depth, semi-structured interviews guided by a protocol detailed in the study’s pre-registration (https://osf.io/nqrk2/). During these interviews, participants were invited to reflect on and map out their experiences of NA in PwD under their care. They were encouraged to provide concrete examples of agitated behaviours observed during the night and to discuss their personal reactions to these events. When participants became visibly emotional, care was taken to ensure they felt comfortable and supported, and they were given the opportunity to pause or discontinue. All participants chose to complete the interview.

### Analysis

This study employed Braun & Clarke’s (2006) six-step framework for thematic analysis. This approach was selected for its flexibility, as it is not bound to any specific epistemological or theoretical stance, making it particularly suitable for exploratory qualitative research (Maguire & Delahunt, 2014). Moreover, since we aimed to gain insight into the experiences of caregivers as well as to map their reactions to the experiences, thematic analysis provided a suitable analytical approach for both aims. For the analysis of transcriptions, we used NVivo 15 software (2020, R1). A bottom-up, inductive approach was adopted, allowing themes to emerge from the data rather than being guided by a top-down method with predefined analytical frameworks (Maguire & Delahunt, 2014).

Before analysis, the first author transcribed all interviews verbatim. This initial phase of transcription also served as the initial phase of familiarization with the data. In the second phase, initial codes were generated using an inductive approach, meaning that coding was not directed by the study’s research questions. Instead, the full transcripts were segmented into meaningful units of data. During the third phase, potential themes were identified. No rigid criteria were applied to define a theme; in some cases, multiple codes were grouped under a single theme, while in others, codes were retained independently for further consideration. The fourth phase involved a collaborative review of preliminary themes by an interdisciplinary team of researchers with a background in psychology, anthropology, philosophy and familiar with qualitative analysis. This process included discussions on the internal coherence of themes and the distinctions between them. Based on this feedback, the first author revised and refined the thematic structure. In the fifth phase, themes were clearly defined and named, resulting in the identification of two overarching themes, each comprising several subthemes. The sixth and final phase, reporting the findings, is represented in the present manuscript. In the following section, we first describe the constituents of the experience of NA from the perspective of informal caregivers, followed by the range of reactions to NA utilized by informal caregivers. We strengthen the trustworthiness of our findings by providing direct quotes from the interviews to illustrate each subtheme. Although the themes are listed separately, we consider them intertwined. For this reason, we put the themes into a conceptual framework in the second part of this section, visualising how the themes operate at different levels – the PwD, the informal caregiver, as well as their environment. Taken together, this constitutes the lived experiences of informal caregivers regarding NA in the persons they care for and their responses to it.

### Researcher reflexivity

As the first author and sole interviewer, I engaged in ongoing reflection on my research position and assumptions about the phenomenon under investigation (Creswell & Miller, 2000). After completing three interviews, I discussed my reflections and experiences with the final author, who provided feedback on the ongoing interviewing process. I do not have personal experience as an informal caregiver of a person with dementia, which enabled me to approach the interviews with openness and attentiveness to participants’ narratives. At the same time, my academic and professional background means I am familiar with the symptomatology of dementia and its potential impact on caregivers, as this is also the focus of my research. I recognize that this knowledge may have influenced the degree of attention I paid to the functioning of the PwD during interviews. To mitigate this, I made a conscious effort to centre my questions and interpretations on the lived experiences of caregivers.

## Results

In line with our research questions, two distinctively important themes emerged from the interviews, namely the experiences of informal caregivers about NA, and their reactions to NA. The theme of caregivers’ lived experiences revealed seven subthemes (sleeping with one eye open, emotional toll of stressful nights, fear for the safety of their loved one, expecting the unexpected, when caring comes at a cost, perceived necessity of medication, external and environmental factors shaping the nights). Regarding the caregivers’ reactions to NA, we identified eight subthemes (emotional reactivity, medication as the last resort, communication as comfort, managing risk through safety solutions, establishing stability through routine, nighttime crisis management, seeking support resources, and learning the hard way).

### Experiences of informal caregivers about nighttime agitation Sleeping with one eye open

Participants described the nights as very challenging, as the circumstances led them to stay alert to what was happening during the night. They explain that “sleeping with one eye open” became an automatic reaction and that, over time, they got used to hearing and recognizing every sound of the PwD. They mention they wake up to every sound and generally recognize what the person is doing by sounds. As they describe that the nights vary, they mention their nights feel like sleeping on “stand by” to notice anything that the night might bring.

> *“I would wake up to everything, I hear everything, I recognize all the sounds, I hear her going to the toilet, walking downstairs…” (Nina)*

### Emotional toll of stressful nights

Interviews revealed a range of emotional experiences of informal caregivers. Caregivers expressed feelings of fear, stress, being lost and worrying about potential falls and aggressive episodes. Fear was often linked to the unpredictability of the episodes and not knowing what could happen. Informal caregivers describe the nights as “hardship” that they cannot hide. Caregivers report being hurt by extremely stressful situations, for example, when PwD become aggressive, and they express not being used to violence. They add that the extent of feeling hurt by such situations also depends on the type of relationship. The emotional toll is significant, with caregivers feeling overwhelmed by the situations at night. Nevertheless, it sometimes not only reflects negative valence, as some caregivers also find moments of feeling at peace and security.

> *“After the diagnosis, the first phase was the most terrifying. I wasn’t used to violence, and my father’s shouting and threats really hurt me. Even though I knew he couldn’t harm me, his aggression overwhelmed me and left me devastated. I felt completely lost and mentally exhausted. My mother kept reminding me that this behavior wasn’t truly him, but the disease.” (Nina)*

### Perceived necessity of medication

Caregivers report that at the beginning, they did not need medication; however, as the disease progressed, they felt the nights became unbearable without medication. They resort to medication to manage challenging situations and ensure they can function the next day. They use phrases such as “I cannot imagine the nights without medication.”, “I have no idea how it would be otherwise.”, “If I don’t give her medication, I cannot go to work the next day.” Still, some caregivers expressed ambivalence regarding medication use. Concerns included administering too much medication, potential dependency on sedatives and the uncertainty about the appropriate timing for consulting a physician to prescribe medication. They also experience concerns about the side effects of the medication and often report that they feel more comfortable with non-pharmacological approaches.

> *“When he broke his hip, the situation worsened, and my mother was also struggling*.

> *Dementia fluctuated between stressful and manageable periods, but eventually, we lost control. At that point, the caregiving burden peaked, so we started with medication. With sedatives, the nights became calmer, and he slept almost all night. Without medication, the situation was unbearable.” (Anna)*

### Fear for the safety of their loved one

Informal caregivers worry about their loved ones leaving the house during the night and not being able to find them. They experience locking the door as very stressful, not only for them but also for PwD. They worry about PwD falling out of bed and are afraid of giving them too much medication for the night when such medication is prescribed only for difficult periods. They experience safety as one of the main issues when it comes to leaving them alone at night, and are afraid of dangerous situations that could happen if they, for example, went away for a weekend. They mentioned they started worrying about unsafe situations once the disease progressed. Only one participant reported that they were not concerned about the safety of the loved one.

> *“The biggest problem is at night…I cannot leave her alone, not even for one night. Someone would need to stay over so that she could be safe.” (Laura)*

### Expecting the unexpected

Caregivers express little difficulty when it comes to predicting that an episode is coming up; however, they struggle with predicting how these episodes would unfold. They explain that they often recognize signs of a challenging night coming up, for example, as the person is acting tired in the evening or is waking up already multiple times in the first part of the night. In terms of predicting the duration of the episode, the reports are ambivalent, as some caregivers express confidence in knowing how long the episode will last, while others describe it as unpredictable. All caregivers express feeling stressed about the unpredictability of the future, not being able to predict the evolution of NA as the disease progresses.

> *“It is all fluid. It comes and goes away like the wind. But the problem is that I never know how long it is going to last and what I can expect if things get worse.” (Kate)*

### When caring comes at a cost

Caregivers also report on their physical and mental well-being as a consequence of agitation at night in the people they are providing care for. Caregivers often experience sleep deprivation, leading to exhaustion and difficulty functioning during the next working day. The strain affects their performance at work, sometimes even with their colleagues noticing changes in their functioning. The constant stress and responsibility are reflected in feelings of guilt, anxiety and emotional breakdowns. Many caregivers express a need for a break or holiday as they feel unable to prioritize their own needs. The burden affects their relationships, family functioning and overall quality of life. Some notice the need to seek therapy themselves, to be able to overcome the challenges and understand better how to respond to the needs and behaviour of their loved ones, and at the same time protect themselves and maintain their private life. Many see themselves on the edge of a burnout.

> *“I need to look after myself. I will soon break under the weight of this. What I would really need is therapy for myself, to learn how to set boundaries and how not to feel guilty in such situations.” (Yasmina)*

### External and environmental factors shaping the nights

Informal caregivers report that agitation during the night can be associated with a variety of factors. They experience fluctuations in NA during the season change, weather change, as well as during the full moon. During the full moon, they observe increased sleeplessness and behaviours, such as wandering. Caregivers also mention that they experience environmental conditions in the bedroom such as the right room temperature and lighting in the room, as highly important for good nights.

Additionally, medical and physical factors such as frequent urination, urinary tract infections, and pain complaints contribute to the difficulty of the nights.

> *“It was also related to his frequent nighttime bathroom visits. On the way back, he became distracted, wandered into other rooms, or got lost and couldn’t find his bedroom. Installing automatic hallway lights helped guide him back to his room and improved the situation.” (Jane)*In the preceding section, we examined how informal caregivers experience NA in PwD. We now turn to how caregivers react and respond to this phenomenon.

### Caregivers’ reactions to nighttime agitation in people living with dementia Establishing stability through routine

Caregivers emphasize that maintaining consistent daily and evening routines is crucial for managing NA. During the day, they find it important to go for a walk so that their loved ones are tired by bedtime. Strategies like calming evening rituals, including warm dinners and a peaceful atmosphere, help the PwD to distinguish between day and night. Caregivers also notice that if the person does not sleep at home (for example, when siblings alternate care responsibilities), the lack of an evening routine contributes to more difficult nights at another place. Caregivers’ descriptions highlight the importance of sleep hygiene for better management of NA.

> *“Once she stayed at my brother’s house while I was away. In the unfamiliar environment, she couldn’t find her way back from the toilet and wandered at night. At home, we have an established routine and an adapted environment, which makes her feel more comfortable. As a result, nighttime difficulties occur less often, and things work much better when she follows a routine.” (Peter)*

### Emotional reactivity

Caregivers’ reactions to emotional hardship regarding the NA often reflected a process of acceptance, recognising that the diagnosis of PwD will not go away and that they must learn to live with it. To not feel hurt or guilty, caregivers try to create distance, especially during challenging episodes. Many work through their thoughts and emotions by discussing them with their friends or a partner, or find comfort in their pets. Others experience helpfulness in continuing to work, as it allows them to “have a reality check”. Support from the partner or other family members allows them to create a system of circling/switching, which lowers their emotional burden and allows them to maintain a positive outlook.

> *“I felt like this was rooted in me. You know it was hard to accept that he became so different*.

> *And now, this doesn’t hurt me anymore. I started to accept it, and it doesn’t hurt me anymore, whatever he tells me. No matter how he is cursing at me. Before, I felt terrible, but now it doesn’t hurt me anymore.” (Nina)*

### Medication as a last resort

Caregivers describe that the difficult situation during the nights was an alarm for them to seek a doctor’s support in terms of starting medication. They mention that introducing medication also established a routine and “made the situation better”. They describe that the solution with medication alleviated the situation for them and the PwD, and that staying on medication allows them to at least have a few nights of good sleep.

> *“It began with convincing but untrue stories. He would leave his room in the middle of the night, moving to another room because he believed he was in a hall listening to a lecture. Often, he was convinced he was not at home but somewhere else, which led him to wander between our two connected apartments, including my sister’s. This usually happened around four in the morning*.

> *These episodes were clear signs that the situation was escalating and raised concerns about potential danger at night. We took it seriously, consulted a doctor, and medication was prescribed for nighttime restlessness.” (Anna)*

### Managing risk through safety solutions

Caregivers often use various safety measures to manage NA and prevent potentially dangerous situations during the night. Many installed cameras or baby monitors to monitor what is happening during the night. Especially in three cases of a dyad, where a caregiver does not share a household with the PwD, they significantly benefit from being able to oversee remotely what is happening during the night. Although many installed cameras, they mention their goal is not to exert control but to provide peace of mind, allowing the caregiver to sleep more calmly. Nevertheless, some caregivers mention that having a camera triggers compulsive checking if the PwD wakes up during the night. All caregivers reported locking the front door during the night to ensure that the person cannot exit the house. Some also considered locking the doors of children’s rooms to avoid nighttime intrusions of the PwD. To ensure safety, some of them use stair fences and bed rails. Next to ensuring safety, such elements also seem to decrease the number of bed exits and wandering around the house.

> *“Often, he would resist or have an outbreak. This happens quite frequently, and he says he is in prison when one of us needs to put up the bed rail when he goes to sleep. He cannot accept that we are putting the bed rail up for his safety, that this is the only way we can make sure nothing bad happens. He would never understand this in the evening, while he always does in the morning. I think it is related to the fact that he is tired in the evening, and it becomes more difficult.” (Anna)*

### Nighttime crisis management

Caregivers frequently described acute situations during the night requiring emergency responses. While some avoided calling 911, others reported taking their loved ones to the emergency room when the situation during the night escalated beyond their control. Some caregivers expressed fear and worry about how to manage future crises and expressed a strong need for clear guidance on whom to call and what steps to take during nighttime emergencies. Caregivers emphasize that during these stressful situations, the PwD often views them as their enemy, which makes the situation unbearable and creates the necessity to be able to call for help.

> *“Sometimes she would hit something or even me, and sometimes my husband needs to intervene as he is stronger than I am. If she is in that moment, she has a feeling that I am her enemy. It depends on the situation, but some situations escalated to the extent that I had to call 911.” (Stephanie)*

### Learning the hard way

Caregivers express frustration about not knowing what to expect regarding the evolution of the situations at night, leading them to rely on experimentation and learning by experience. They mention “being a layman” in this, which forced them to learn through experience and respond to what “felt like the right approach”. They acknowledge that they cannot prevent every situation, but they emphasized a strong desire for clearer guidelines and proactive warnings from professionals on how to handle difficult situations during the night.

> *“I tried to handle everything related to my mother’s dementia as calmly as possible, but sometimes it became too overwhelming. Especially without any professional help at home. Because formal caregivers are somehow the ones who know how to do things, they can offer reliable assistance and support. We, informal caregivers, are lay people; we act based on our feelings, but we learn by experience. You hope you have done something good for her.” (Yasmina)*

### Communication as comfort

Caregivers emphasize that while managing NA, good communication plays an important role. As a first solution to an episode of agitation during the night, they mention calming down the PwD, using a soft voice, gentle gestures, and slow, understanding speech with open-format questions. They also mention the type of calming down is different from one situation to another. Despite acknowledging that communication is crucial in the management of NA, caregivers complain that it often takes a lot of verbal persuasion and negotiation for the PwD to go back to bed. In such cases, they are aware that their communication can be rougher, which in turn also reinforces resistant behaviour in PwD in that moment. This reflects the dual role of communication in managing NA. Despite the caregivers using it as a comfort strategy, when used inappropriately, it can also be viewed as a potential source of tension or a trigger for agitation.

> *“In the past, I sometimes had to physically move her away from the door because I didn’t know what she might do next. She would start screaming and calling. Now, I leave her there and try not to intervene, as that would make her more tense. Instead, I distract her with open questions like, “Mom, what do you see? What happened there?” This redirects her attention and helps her calm down.” (Yasmina)*

### Seeking support services

Caregivers seek various resource options to manage the nights effectively; however, they also express a lack of certain support tools/services. Many found valuable advice through spoken interactions with other caregivers on social media platforms or by searching for answers in dementia-related books.

While some caregivers opt for finding information online, others value personal contact and assistance from the medical field. Although some caregivers took courses to improve their caregiving skills, they still highlight the need for more targeted support groups and resources focused on the late stages of dementia and particularly challenging symptoms such as NA, which require more advanced knowledge and skills for their management. A recurrent concern was the lack of information about what to expect in the future as the disease progresses, leaving caregivers with doubts about when and how to seek assistance in difficult situations at night.

> *“I prefer personal contact, so I decided to have a consultation as a plan B for emergencies, to help me stay calm. I met a social worker who specializes in dementia and has good contacts with nursing homes. I shared my moral conflict about transferring my mother, but needed support and to know there are options if her behavior worsens. If she were to wake me every night, escalate to hallucinations, or become dangerous, I couldn’t manage. Knowing a support system exists helps me handle risks, maintain my responsibilities, and feel calmer about the future.” (Laura)*

### Layered perspectives of informal caregivers

The layered perspectives overview was created with the purpose of presenting the results in a more accessible and contextualized manner. As presented in *Figure 1*, experiences as well as reactions to NA reported by informal caregivers span over three levels, namely, the level of the PwD, the level of the caregiver, and the level of the environment. Participants’ stories were characterized by the interplay of all three levels, which reflects the complexity of the phenomenon. By zooming in, we can observe that despite the spread at different levels, most of the phenomena occur within the PwD-caregiver dyad. This suggests that the informal caregiver – PwD dyad captures the essence of experiences and reactions to NA, showcasing that mitigating this phenomenon with a perspective of the dyadic relationship might offer a crucial opportunity for targeting interventions. The dyadic relationship was in many cases, put into the context of the environment, where caregivers, despite striving for, viewed lack of support and guidance from the environment and the systemic/professional side as very important. Participants’ stories reflect the expression of the need to not only contextualize NA at the level of the PwD, but also utilize a more holistic approach and not forget about the importance of the caregiver and the environment when talking about the phenomenon of NA. This has also been seen in the observation that while informal caregivers were interviewed about the NA in PwD, all of them extensively reflected upon their mental and physical well-being as a consequence of the difficult nights. Hearing caregivers talk about their well-being reminds us of the importance of recognizing the need for support for the informal caregivers, who, despite trying out different strategies to address NA in PwD, continue to feel exhausted.

**Figure 1.**
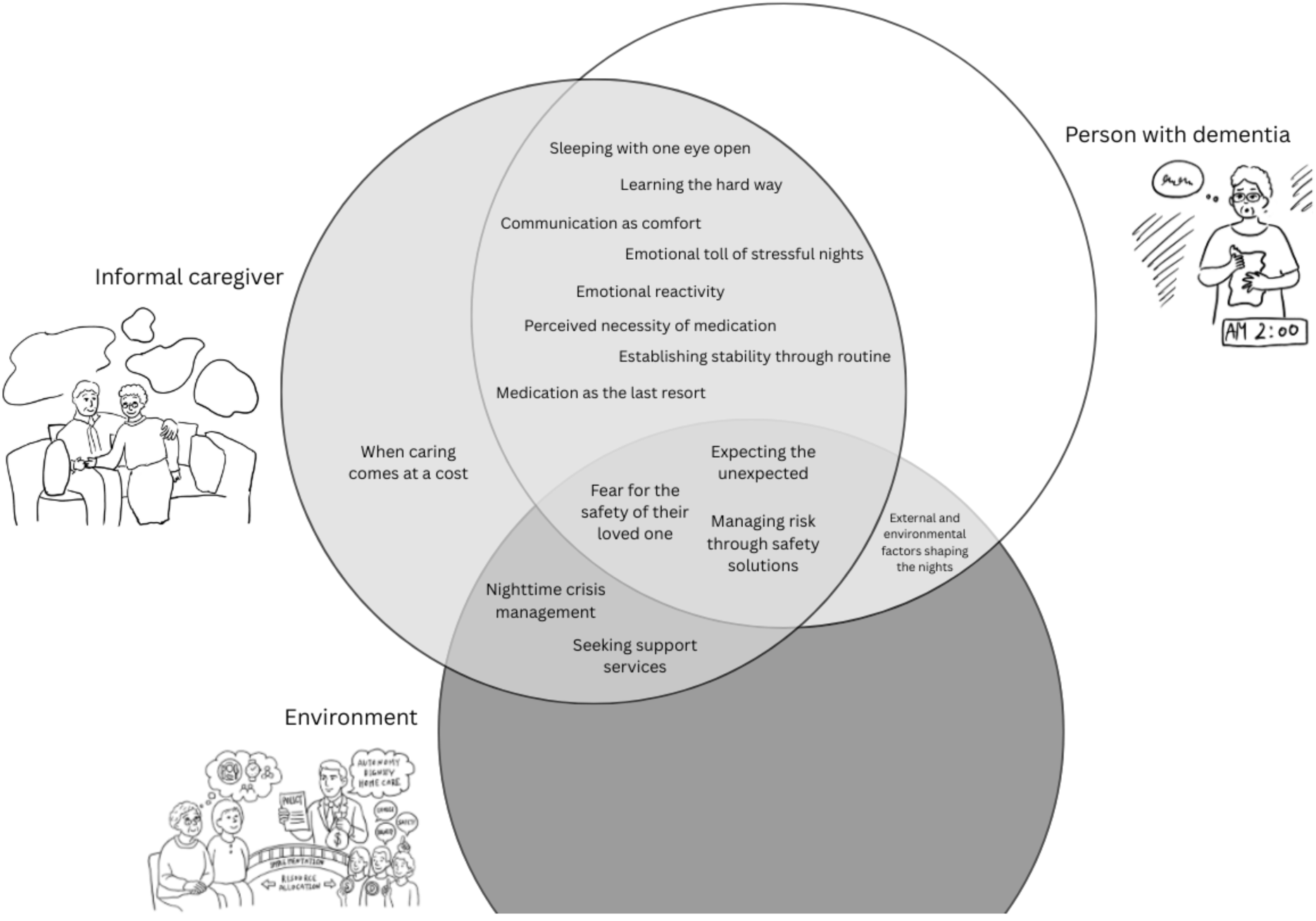
Layered perspectives of informal caregivers regarding NA

## Discussion

With this study, we aimed to address the lived experiences of informal caregivers of PwD on NA and explore their reactions to this phenomenon. Our results revealed the complexity of NA and show that it is not experienced solely as an individual symptom of PwD but rather as a multifaceted phenomenon unfolding across three interconnected levels: PwD, informal caregiver, and the environment. The majority of identified experiences and reactions were identified at the dyadic level, highlighting the interdependent nature of NA and its management within the caregiving relationship. We noticed differences in caregivers’ experiences depending on the nature of nighttime disturbances in PwD. When PwD simply wake up during the night, caregivers typically react differently than when those awakenings are accompanied by NA, such as wandering or resistance to return to bed. These episodes can escalate into moments of aggression or, if unnoticed by the caregiver, lead to potentially dangerous situations, such as the person leaving the house or engaging in activities inappropriate for nighttime. As a result, caregivers described a constant need to remain alert throughout the night, reflecting the unique challenges posed by NA compared to sleep disturbances alone. Across interviews, caregivers often shifted their focus from describing the experiences with PwD to emphasizing the toll NA took on their own mental and physical well-being. This narrative shift highlights that NA is not only a behavioral or clinical concern for PwD, but also a significant stressor for caregivers, impacting their health, coping capacity, and sustainability in the caregiving role. Our results emphasize the need for interventions that address both symptom management in PwD and the well-being of informal caregivers.

Two overarching themes, caregivers’ experiences and reactions, emerged with seven and eight sub-themes, respectively. One of the most common experiential themes was captured by the metaphor “sleeping with one eye open”, a phrase already identified in the literature on dementia caregiving and nighttime disturbances (Huisman et al., 2025; Spring et al., 2009). Caregivers described the nights as characterized by constant vigilance, fragmented sleep, and heightened anxiety, driven by the anticipation of potential agitation or unsafe incidents. Sustained alertness prevented caregivers from obtaining a sufficient amount of sleep and contributed to cumulative exhaustion, reinforcing their poor mental and physical well-being.

The emotional consequences of prolonged nighttime vigilance were evident. Caregivers reported experiencing anxiety, irritability, guilt, low mood, and feelings of hopelessness, reflecting previous findings that link sleep disturbances in PwD to caregiver emotional distress (Spring et al., 2009). Our findings extend this literature by showing that NA, in particular, can evoke complex and emotionally challenging reactions. For example, when PwD displayed aggression during nighttime episodes, caregivers often felt emotionally hurt, especially when such behaviors were inconsistent with the person’s personality before dementia’s onset. These experiences reflected feelings of grief and loss and contributed to emotional strain beyond that associated with sleep disturbances alone.

Over time, the cumulative effect of chronic sleep deprivation, emotional burden, and sustained vigilance had a detrimental impact on caregivers’ overall well-being, consistent with prior research (Gibson et al., 2023; Gibson et al., 2014).

One of the key drivers for caregivers’ nighttime vigilance was concern for the safety of PwD. Fear for the safety of a loved one during the night has been consistently reported in studies of dementia-related sleep disturbances (Gibson et al., 2014; Greenblum & Rowe, 2012; Huisman et al., 2025; Spring et al., 2009), and our study shows that NA intensifies these concerns. Caregivers described fears related to wandering, falls, aggression, and unsafe behaviors resulting from disorientation. When agitation co-occurred with nighttime awakenings, caregivers perceived nights as significantly more challenging due to the unpredictability of PwD’s behavior and the risk of losing track of the person. In response to these safety concerns, caregivers adopted a range of management strategies, including bed rails, locking doors, and relying on monitoring devices. The use of such strategies often varied depending on disease progression and the caregivers’ perceived balance between safety and autonomy. While these measures, for example, monitoring devices, were intended to mitigate risk, they frequently reinforced caregivers’ sense of responsibility by constant monitoring, further disrupting caregivers’ sleep. This suggests the potential value of supportive technology that can reliably detect changes in PwD’s nighttime behavior and alert caregivers only when intervention or check-up is necessary, thereby reducing continuous hypervigilance and allowing caregivers to rest more efficiently (Spring et al., 2009).

### Supporting informal caregivers using a dyadic approach

A central contribution of this study is the identification of NA as a dyadic phenomenon. As most of the identified experiences and reactions involved both PwD and their informal caregivers, our findings emphasize the importance of addressing NA as a relational process rather than solely as an individual symptom. Despite previous research demonstrating the relevance of dyadic coping in dementia care, most studies continue to focus on either PwD or caregivers in isolation, thereby overlooking relational dynamics (Braun et al., 2009).

Our study extends dyadic coping theory by illustrating how NA functions as a context-specific stressor that is jointly experienced, interpreted, and managed within the caregiving dyad.

As shown in *Figure 1*, responses to NA are interdependent, with caregivers’ actions influencing PwD’s behaviors and vice versa, highlighting reciprocal processes of adaptation and regulation. Dyadic coping theory proposes that stressors related to chronic illness are jointly appraised and managed, with consequences for both partners’ well-being (Braun et al., 2009), and previous work in dementia has shown that dyadic coping mediates the relationship between caregiving stress and caregiver quality of life (Häusler et al., 2016).

Consistent with emerging evidence from dyadic sleep interventions in dementia, which suggests benefits for both PwD and caregivers when both members are involved in nighttime management (Song et al., 2023), our results support the development of interventions that target dyadic functioning. Such interventions should be tailored to dyad-specific characteristics, recognizing that caregiving relationships differ in communication patterns, coping styles, and risk tolerance.

Importantly, caregiver well-being should be positioned as a central outcome of these interventions, rather than as a secondary or indirect benefit.

### Clinical implications and targets for interventions

The next step is to attempt to develop interventions to manage NA while taking into account the needs of informal caregivers identified in our study. Despite the significant clinical impact of NA, its management remains challenging, due to its heterogeneous nature and the wide range of potential triggers (Van Den Bossche & Vandenbulcke, 2021). Our findings align with existing literature identifying both medical factors (e.g., pain, urinary tract infections, medication side effects) and environmental factors (e.g., overstimulation, inadequate lighting, nighttime noise) as contributors to NA (Richards et al., 2021; Rose et al., 2015). Notably, caregivers emphasized that these triggers can vary not only between individuals but also within the same person over time, further complicating management. Caregivers consistently expressed a preference for non-pharmacological interventions for NA. However, many reported feeling compelled to resort to antipsychotic or hypnotic medications when situations became overwhelming. Yet, there has been no consistent evidence for the effect of such medications on NA specifically, and their use is often accompanied by adverse effects (Ballard et al., 2009; Rosenheck et al., 2007).

At present, no standardized interventions specifically address NA in PwD. As a result, caregivers described being forced to “learn the hard way”, relying on trial-and-error approaches and self-initiated strategies to manage difficult nighttime behaviors. While standardized non-pharmacological interventions exist for daytime agitation (Kong et al., 2009; Livingston et al., 2014) and sleep disturbances (Cipriani et al., 2015), these interventions are not designed to address NA as a distinct phenomenon. Although NA overlaps with both daytime agitation and sleep disturbances, it presents unique challenges related to the nighttime context, including heightened caregiver burden, reduced access to support, and different triggers such as nighttime awakenings or insomnia (Flisar et al., in preparation). Consequently, interventions targeting agitation or sleep problems in isolation are insufficient when these issues co-occur at night. Integration of sleep-focused and NA-focused interventions is crucial as these phenomena frequently reinforce one another during the night (Rose et al., 2011). An example of a promising framework is DREAMS-START, a multicomponent, personalized-based intervention designed to support caregivers and PwD with sleep disturbances (Livingston et al., 2019; Rapaport et al., 2018). Although DREAMS-START primarily targets sleep disturbances, it includes components addressing difficult nighttime behaviors and aligns several themes identified in our study, such as establishing routines, managing environmental factors, and providing psychoeducation. While NA is not explicitly labelled as such within this intervention, it represents an important step toward addressing the complexity of nighttime caregiving.

Our findings suggest that NA may require more advanced skills and targeted support than sleep disturbances alone. Caregivers may benefit from training in communication strategies, anticipation of NA episodes, safety planning, and crisis management, as well as guidance on balancing safety with autonomy. To achieve this, building on existing intervention frameworks while incorporating the specific challenges for NA offers a promising direction for future research. For example, to support crisis management, we propose a traffic-light–based signalling system to help caregivers judge the severity of nighttime situations and determine appropriate responses based on safety risks and caregiver strain, with tailored coping strategies linked to each level of urgency (e.g., green, orange, red).

Based on our results, we recommend the following. First, future studies should stratify support according to nighttime symptom severity, recognizing the distinct caregiving needs associated with sleep disturbances alone versus NA. Second, caregiver training should specifically target NA and include skills for managing agitation, aggression, and unsafe/challenging nighttime behaviors. Third, caregivers should have access to emergency planning and rapid-response support for high-risk nighttime situations. Fourth, caregiver-focused mental health support, such as counselling, should be integrated to address boundary-setting and reduce caregiver guilt. Finally, caregiver well-being should be recognized as a core outcome, rather than a secondary consideration, in the development of dementia care interventions.

## Limitations

This study should be considered in light of several limitations. Interviews were conducted in Slovenia, a familialist context in which intergenerational caregiving is common and strong cultural expectations regarding family care may contribute to higher caregiver burden, guilt, and worry (Filipovic Hrast et al., 2020). As such, the findings are context-specific and may not be fully generalisable to settings with different care arrangements. Additionally, analyses were based on English translations to facilitate team discussion, which may have resulted in the loss of linguistic nuance present in the original Slovenian interviews. Given the qualitative design, the researcher subjectivity may have influenced data interpretation; this includes the first author’s cultural background and the age difference between the researcher and participants, which may have shaped the understanding of caregivers’ experiences.

## Conclusion

Given the high reliance on informal caregivers in dementia care, understanding their perspectives on challenging symptoms in dementia is crucial. The current study identified a wide range of caregivers’ experiences and reactions to NA in PwD. Interventions should be focused on three identified levels at which the phenomena occur, namely, PwD, informal caregiver and the environment. With most of the themes occurring at the dyadic level of PwD and the informal caregiver, future interventions should target this interaction sufficiently. Interventions to address sleep disturbances and agitation in general exist; however, they insufficiently address the needs expressed through the lived experiences of carers identified in our study. We suggest that sleep and agitation-focused interventions should be integrated to provide more advanced support and skills for NA, which is more challenging when it is intertwined with sleep disturbances. Lastly, informal caregivers devoted a significant focus to deterioration of their own mental and physical well-being as a consequence of NA, highlighting that when developing interventions, maintaining carers’ well-being is equally important as managing NA in PwD.

## Data Availability

All data produced in the present work are contained in the manuscript

## Footnotes

1 This study was part of a larger study, in the current paper, we only present the results on the experiences and reactions of informal caregivers to nighttime agitation in people living with dementia. The reason for this was the extend of the collected data.

